# Crimean-Congo haemorrhagic fever virus transmission: exploring perceptions of human-animal-tick interactions across six districts in Uganda

**DOI:** 10.64898/2026.06.10.26355422

**Authors:** Marina Kugler, Lazaaro Mujumbusi, Lucy Pickering, Richard Muhumuza, Mathias Akugizibwe, Edward Obicho, Titus Apangu, Evalyne Umo, Simpson Nuwamanya, Shirin Ashraf, Stella A Atim, Emma C Thomson, Poppy H L Lamberton

## Abstract

Crimean-Congo haemorrhagic fever virus (CCHFV) causes a viral zoonotic disease transmitted through tick bites and direct contact with infected blood or tissue of infected animals. Socio-ecological and behavioural risk factors for CCHFV exposure in Uganda remain poorly understood, which can lead to the omission of key risk factors in quantitative survey design and limit our wider understanding. In this study, we explored human-animal-tick interaction transmission risks in Uganda.

We conducted 24 focus group discussions (FGDs) and 31 key-informant interviews (KIIs) across six environmentally and socio-ecologically diverse districts, between October 2023 and March 2024. Study sites were selected using K-prototype analysis, which combined environmental and socio-ecological variables to identify distinct clusters within Uganda. FGDs were conducted separately with groups of community leaders, men, women and teenagers with stratified purposive sampling. Medical doctors, veterinarians, traditional healers, district surveillance officers, and herdsmen were individually interviewed as key informants and purposively sampled. Data were transcribed and translated into English, and analysed thematically using iterative categorisation in NVivo 14.

Most participants reported tick bites, some as frequently as every day. Close contact with animals was common, including sleeping next to them in the same building, largely due to concerns about animal theft. Less frequent but notable practices included slaughtering animals for consumption or sacrifice and interactions with wild animals during hunting. Slaughtering and butchering an animal which was sick or had died was reportedly performed by participants in most districts. Plucking and roasting engorged ticks was a practice described in the Kaabong and Arua districts of Northern Uganda.

These practices and behaviours highlight potential key risks of CCHFV transmission and underscore the need for future studies to address specific behaviours, to quantify if, and to what extent, they present an exposure risk. Further work should include underlying reasons for the behaviours, which would help ensure that culturally appropriate interventions are targeted.

## Introduction

Crimean-Congo haemorrhagic fever virus (CCHFV) is a zoonotic virus that can infect domestic and wild animals and is thought to be mainly spread indirectly by tick bites, as well as directly, through contact with infected animals or other human cases (1,2). Most human infections with CCHFV result in a mild, non-specific febrile illness (3). Bodur et al. estimated in their study in Turkey that 88% of cases there are subclinical (4), and modelling efforts of infections in Afghanistan suggested that 69% of cases there are subclinical (5). However, some patients develop a severe haemorrhagic disease, characterised by bleeding into the skin, bleeding from the gastrointestinal and urinary tracts, hepatomegaly and splenomegaly, which can lead to death (3). Case fatality rates vary (6), with a study reporting 31.2% mortality in patients hospitalised with CCHFV in Uganda between 2013 and 2019 (7). Understanding transmission risks by exploring interactions of humans with ticks and animals is essential for delineating risk factors for exposure to CCHFV.

Previous risk analyses evaluating exposure to CCHFV with variables from the literature (8–10) include common behaviours, such as caring for animals, slaughtering animals, and hunting. However, specific behaviours and activities regarding these activities and interactions with animals, ticks and animal products vary throughout the world, and may even be different in neighbouring communities. Multiple surveys have investigated risk factors for CCHFV infections and exposure in Uganda (7,8,11). However, only Atim et al. included the practice of eating engorged ticks in their survey, which emerged as a significant risk factor for exposure to CCHFV (8). Eating engorged ticks is not limited to Uganda: A recent study in Cameroon reported that 3% of interviewees had eaten ticks (12). Further factors are also likely to be under-reported and under-investigated by researchers and may be key to better understanding exposure risk and identifying opportunities for intervention.

Few qualitative studies have been conducted regarding CCHFV and other tick-borne diseases. Most qualitative studies around tick-borne diseases have been carried out in the Northern Hemisphere with a focus on Lyme disease (13–15). Qualitative studies focusing on CCHFV have included surveyed knowledge, attitudes and practices (KAP) questionnaires in Pakistan (16) and Burkina Faso (17), as well as a One Health joint risk assessment in Nigeria (18) with a similar approach in Bhutan (19). The first qualitative study on CCHFV in Uganda was published in 2023 by Ayebare et al., and focused on knowledge, attitudes, and control measures (20). In this study, participants mentioned that they slaughtered animals at home and that some ate half-cooked meat. This study highlighted useful new findings (20). However, it was only conducted in Kagadi district of Uganda (Western Region), and did not address other areas in Uganda, nor any comparisons on potential risk behaviours across areas. Since then, Agaba et al. (2025) explored several zoonoses, including CCHFV, in Uganda. They noted the importance of regional differences, noting different exposure pathways in areas bordering national parks, as well as highlighting the importance of human gender roles in animal husbandry (21).

Countrywide and comparative studies across regions, environmentally distinct areas, or tribes on potential CCHFV exposure risk behaviour are lacking in Uganda and could improve current understanding and mitigation strategies against CCHFV transmission. Qualitative studies can aid in gaining a deeper understanding of behavioural risk activities by asking open-ended questions and listening to participants’ stories without predetermined ideas. This is the first study comparing environmental and socio-ecological distinct districts in Uganda using focus group discussions (FGDs) and key informant interviews (KIIs) to understand participants’ perspectives on animal-human-tick interactions.

The overarching aim was to highlight potentially relevant transmission routes of CCHFV through ticks and direct contact with infected animal products and whether these vary across distinct environmentally and socio-ecological settings in Uganda. This aim was addressed through FGDs and KIIs focusing on four key objectives: (1) To identify the perceived burden of ticks and tick bites in each district and to highlight differences in tick bite risk across environmentally and socio-ecological distinct districts in Uganda. (2) To characterise interactions with ticks which could potentially expose humans to CCHFV. (3) To analyse and understand slaughtering locations and methods, to understand the potential risk of direct transmission. (4) To identify differences between animal product consumption practices to identify potential risk of transmission.

## Methods

### Study sites selection

Previous research on CCHFV highlights the importance of study location in determining exposure risks in Uganda (8). These risks may stem not only from environmental differences, such as rainfall, temperature, or land cover, but also from socio-ecological factors, which describe how humans interact with their surroundings. To identify districts within distinct environmental and socio-ecological clusters in Uganda, we employed K-prototype analysis (22) using the clusterMixType library (23) and incorporated 20 environmental and socio-ecological variables. Sources for all variables and example figures plotted on a map of Uganda are presented in S1. Numerical variables were scaled (24), and the elbow method, which identifies the point at which the decrease in the total within-cluster sum of squares slows (25), was used to determine the optimal number of clusters. 13 distinct clusters were identified. Of these, six were selected for the study based on previous work in the district to facilitate logistics, while also considering the largest differences across the variables used (**Figure 1)**.

**Fig 1.**
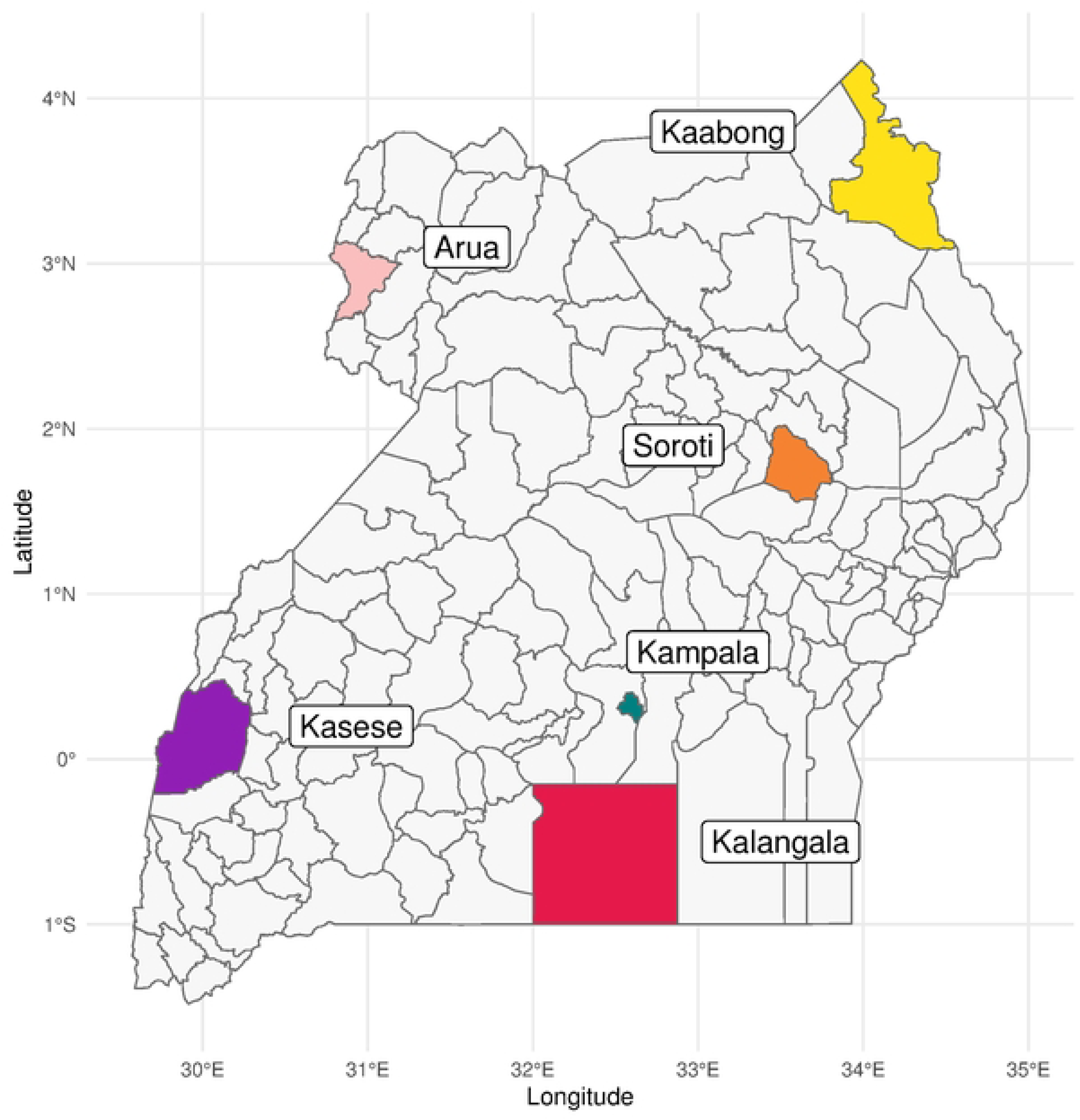
Six selected study districts within Uganda. Selected through initial K-prototype analysis based on 20 environmental and socio-ecological variables and reduced to the selected six districts for ease of logistics and the largest differences within the variables used.

### Study sites description

The six study districts vary substantially in their environmental and socio-ecological variables (S1 Figure 1). **Table 1** presents each selected district, its population size, land area, estimated per capita gross domestic product (GDP), and climate classification by Koeppen (26–30).

**Table 1.** District population, land area, estimated GDP per capita and climate classification. Population numbers are taken from (26). Land area is recorded from Wikipedia entries (27), and shows additional total areas for Kalangala (the land area is much smaller than the total area, as they are islands in a large area of Lake Victoria), and Kasese (contains two large National parks which are not used by its human population). The estimated GDP per capita is taken from (28), and the climate classification is the Köppen classification (29,30).

| District | Region | Population | Land area<br>(total area)<br>[sq mi] | GDP<br>per capita | Classified<br>climate |
| --- | --- | --- | --- | --- | --- |
| Kampala | Central | 1,800,000 | 73 | \$ 2,655 | Tropical rainforest |
| Kalangala | Central | 74,000 | 180.8<br>(3,514.7) | \$ 67 | Tropical monsoon |
| Kasese | Western | 854,000 | 458<br>(1,052) | \$ 540 | Tropical savanna |
| Arua | North | 160,000 | 1,249.6 | \$ 261 | Tropical savanna |
| Soroti | East | 266,000 | 545.1 | \$ 586 | Tropical savanna |
| Kaabong | North | 265,000 | 2,789.1 | \$ 75 | Tropical savanna |

### Study design and data generation

This was a cross-sectional qualitative study conducted in six environmentally and socio-ecologically distinct districts of Uganda. We conducted a total of 24 FGDs and 31 KIIs across the six districts, between October 2023 and March 2024.

This included four FGDs in each district with the following characteristics:

- a group of community leaders, including local councils, religious leaders, and women’s leaders
- a group of men between 18 and 30 years old (as opposed to rather older men usually presented in the community leaders)
- a group of women between 30 and 60 years old
- a group of teenagers, with mixed sex and varied educational levels.

All FGDs sites were selected to maximise geographic, urban/(peri-urban)/rural gradients, socio-ecological and environmental differences, where financially and logistically feasible. An exception was Kampala, the capital and largest city of Uganda, where we focused on only one of the five divisions rather than the entire city. We chose Kawempe division, selected deliberately as a very typical division representing Kampala as a whole, based on local knowledge, at best. The convenient selection of FGD participants in all districts was undertaken by mobilisers within the village centres, after explaining the study aims and the broad categories into which participants should be categorised, including age, sex, occupation and animal ownership.

Five KIIs were conducted in each district, with purposively selected participants: a medical doctor, a veterinarian, the district surveillance focal person (public health specialist overseeing all public health surveillance activities for the district), a herdsman, and a traditional healer or herbalist. In Kampala, an extra KII was added with a meat inspector.

This study was coordinated from the University of Glasgow and conducted in Uganda in close collaboration with in-country researchers, who led data collection within the districts and contributed to analysis and interpretation. Due to the different local languages in each district, six social scientists, who spoke the representative languages and English, conducted the KIIs and guided the FGDs, each in the languages preferred by the study participants (**Table 2**). Many KIIs were conducted in English at the participants’ preference, and in these cases, MK interviewed the participants. All others were conducted as highlighted in **Table 2**. We recognise that the researchers’ positionality, including their institutional location, their social position, sex, and proximity to the study context, shapes the research process. Reflexivity was addressed through ongoing discussions within the research team to support transparency.

**Table 2.**
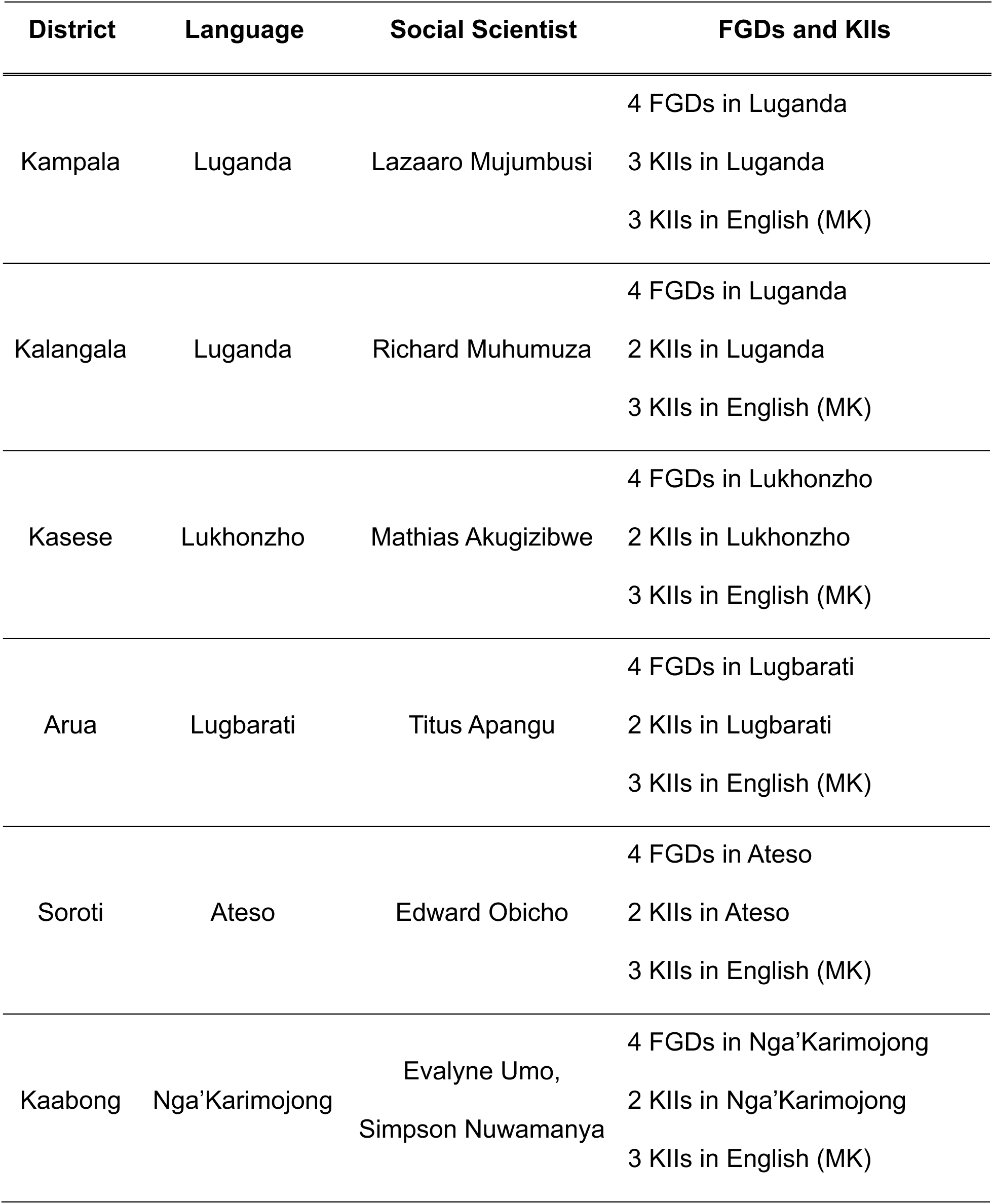
Social scientists working in the six districts. For each district, we had a social scientist who spoke the local language as their mother tongue. The table shows the districts with their most common local language spoken, the social scientist conducting all FGDs and KIIs in the language and the numbers of KIIs conducted in English by MK.

Demographic data of all participants in the FGDs were recorded in REDCap (31) (see S2). To guide the FGDs and KIIs, semi-structured topic guides were created (see S2), including a variety of questions related to animal husbandry, tick interactions and perceptions about transmission risks through tick bites and direct contact. We also discussed recommendations and comments, and the participants were invited to ask questions throughout the sessions.

All FGDs and KIIs were recorded on a digital recorder and translated to English and transcribed by the respective social scientists, as shown in **Table 2**.

### Data analysis

To analyse the data, firstly, four transcripts (women’s FGD Arua, community leaders’ FGD Kalangala, one KII from Kampala, and men’s FGD Kaabong) were coded inductively by MK and LM, to ensure that practices, understandings and perceptions of transmission risk unanticipated by the review of literature were included. MK and LM compared their coding and created a single code book with parents and child codes and detailed explanations for each code. ‘Other’ options were added to ensure that unanticipated practices could be included in the further coding.

All further coding was conducted using NVivo Version 14 (32). Independent coding was performed by three researchers, RM, MA, and MK, and were combined into a single project on NVivo and extracted using R (33) into a Microsoft Excel document (34). Iterative categorisation (IC) was used to analyse the child codes related to the possible risk of transmission to CCHFV, to generate themes within codes and to prioritise themes to present as output (35).

### Ethical considerations and permissions

The study falls under a wider project, approved by the UVRI REC (GC/127/18/09/662) and by the UNCST (HS 2485). An amendment containing the details of this study was approved on the 7^th^ February 2023. The study was introduced and explained to the district health officer (DHO) and other district officials, where appropriate, and approval was sought to conduct the study within the district and local communities prior to any data generation.

Informed written consent was obtained from all study participants. An information sheet in the local languages, containing the necessary details about the study, ethics, and data usage was shared, read aloud, and discussed in the group, giving people time to ask questions and have them addressed. All forms were available in English and the district’s main local language. To signal consent to take part, participants were asked to either sign or provide a witnessed thumbprint on the consent form (see S2). For teenagers, a parent/legal guardian signed the official parental/legal guardian consent form document, which was either explained and read to them the day before by the local mobiliser, or the parent/legal guardian joined on the day of the FGD and left after the signatures took place. All participants in the teenager FDGs signed or provided a thumbprint on an assent form.

## Results

### Characteristics of study sites and study population

A total of 152 participants took part in the 24 FGDs and 31 KIIs across the six study sites. All demographics are presented in **Table 3**. Overall, more men participated than women (61.2% vs 38.8%), and most participants were Christians by religion with few Muslims (11.2%). Most participants in the KIIs studied at universities (including medical doctors, or veterinarians), but some have no or basic education (traditional healers, or herdsmen).

**Table 3.**
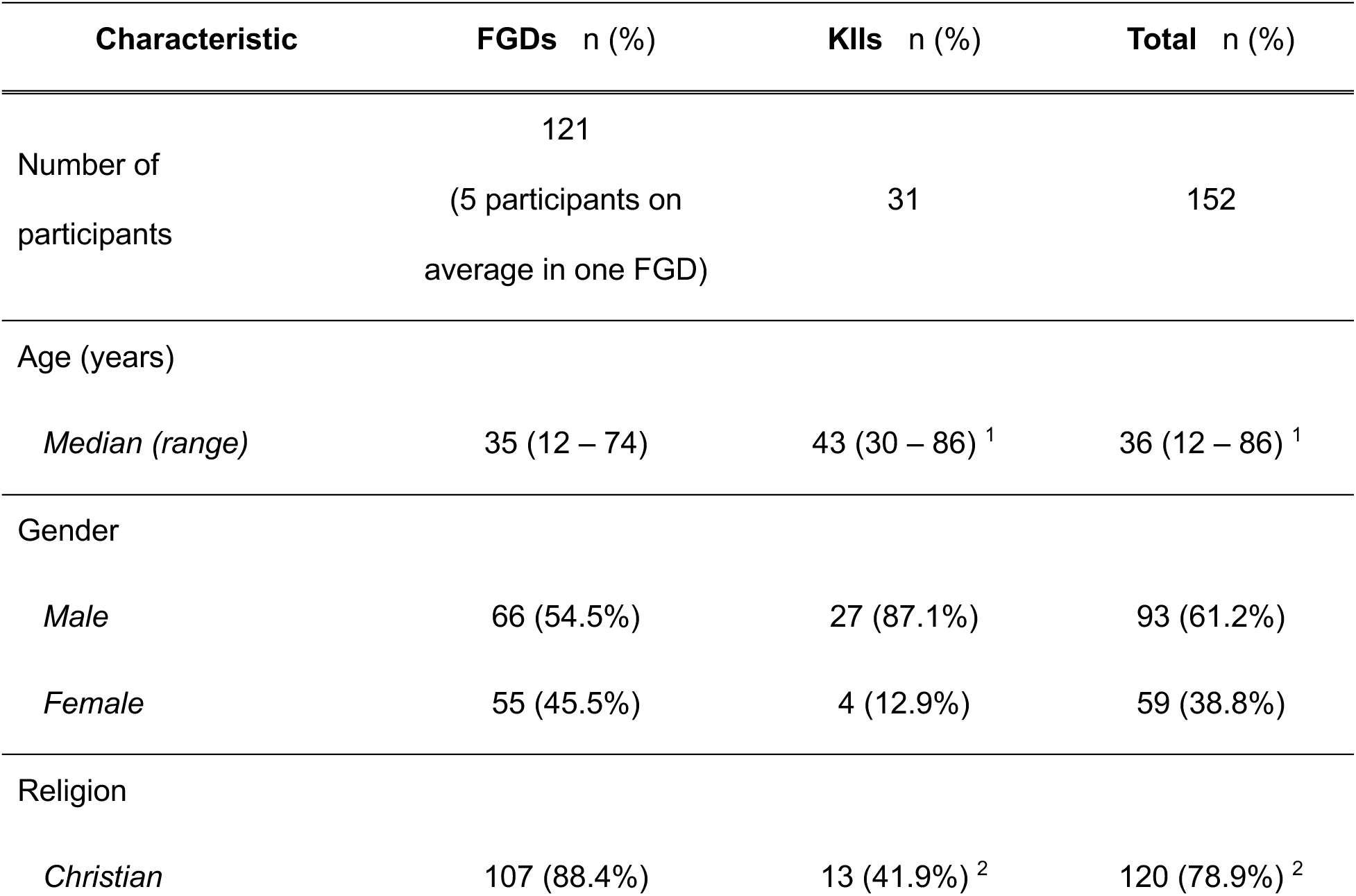
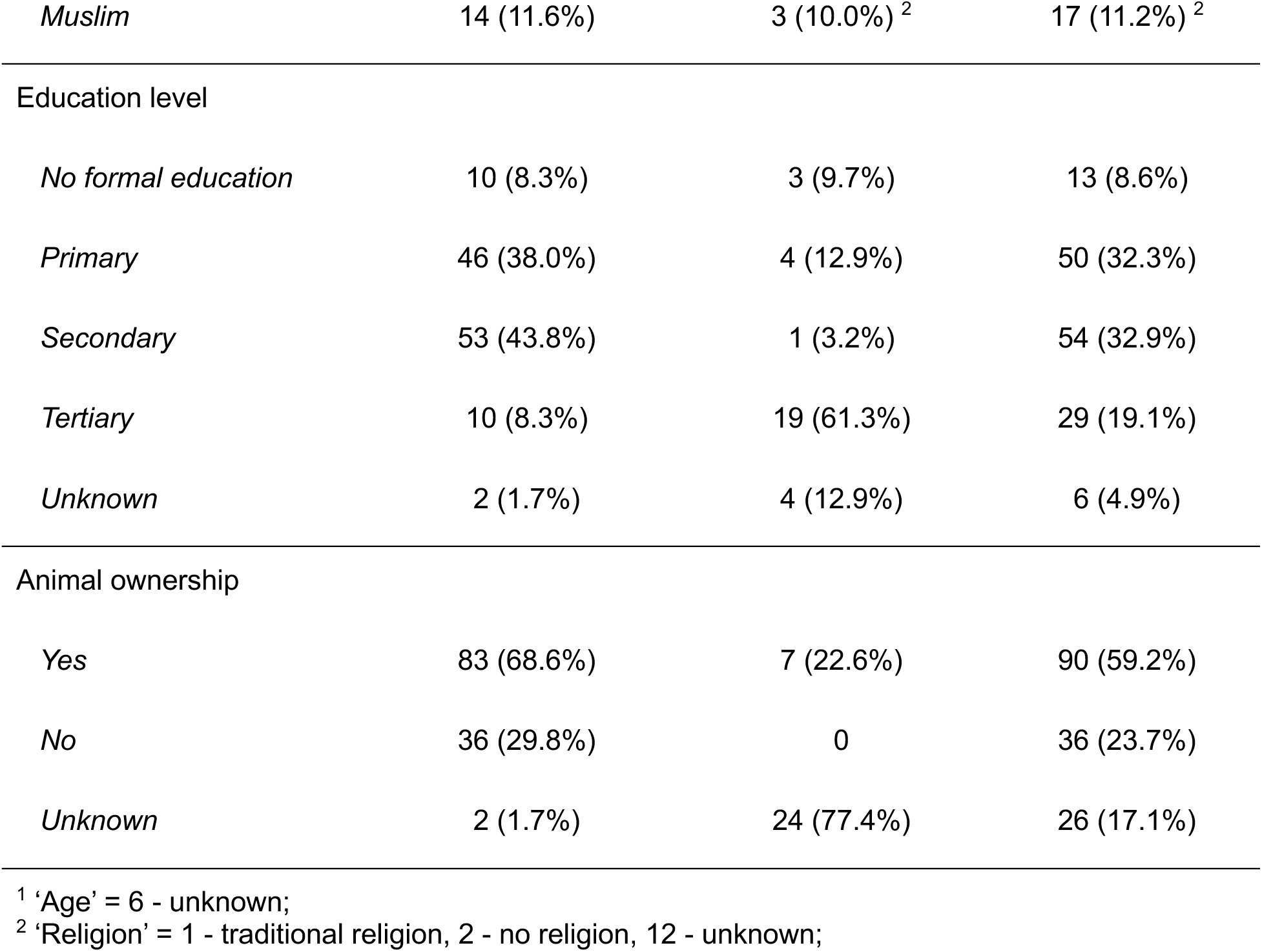
Demographics of the study population. During the KIIs and FGDs, participants described the study districts, and the reports can help contextualise the study within Uganda. Detailed descriptions and quotes are presented in S3, including photographs of the study sites.

### Analysing different transmission routes for CCHFV

In the FGDs and KIIs, the moderator guided through the general transmission routes for CCHFV, and following this, different practices and events that could put people at risk of CCHFV infection in their district were described. Several aspects were discussed, and four key categories arose from the analyses. These were discussed at varying points throughout the FGDs and KIIs, but are presented here separately, ordered by the likely importance that such a behaviour might have on CCHFV infection risk, based on literature knowledge. These four main categories were: risk of tick bites, tick consumption, direct contact during slaughtering, and animal product consumption.

### Risk of tick bites

Tick bites were reported by participants in all districts. However, they were reported more commonly in some districts (Kaabong, Soroti, Arua, and Kasese) than others (Kalangala and Kampala). Participants in Soroti and Kasese reported the most bites, with many reporting to have been bitten many times.

> *“[Burst into laughter] It [tick bite] is a common thing in our community.”* (Men’s FGD, Soroti)

The abundance of ticks described by the participants tended to be reflected by the number of reported tick bites within the community, including in the environment and on domestic animals. A notable situation happened while conducting the men’s FGD in Kasese district, where the participants found ticks just below them in the grass. Three ticks were found in ankle-high grass under a tree shade during the FGD.

> *“Actually, you may find a tick here because goats do graze here [points to the grass]. Yeah, it’s even here.”* (Men’s FGD, Kasese)

This observation highlighted the most common answer to where ticks were found in the environment, which was where animals graze, feed, drink, or rest. Participants also described commonly seeing ticks in high grasses and bushes. Additionally, in Kaabong and Kasese, both areas with large national parks and wild animals nearby, participants mentioned that ticks are commonly seen there and that there is a risk of getting tick bites when entering parks to collect firewood or to poach wild animals for meat. These were also reported to present a risk to domestic animals, which share water sources with wild animals.

> *“Now when we take the cows or goats for grazing, you find that the ticks have already been dropped there by these wild animals, so [our] animals end up bringing ticks back home.”* (Community leader’s FGD, Kaabong)

Across all districts, a view shared by most participants, was that children and young adults are most susceptible to getting bitten by ticks. Multiple reasons were stated as to why children and young adults are considered particularly susceptible, and the majority related to their contact with animals, and playing in the grass. Also many older people reported to have been bitten by ticks a lot when they were young, while looking after animals.

> *“The young ones that play around the compound, playing with the dogs and goats, [are most susceptible to tick bites.]“* (Women’s FGD, Kalangala)

Kalangala, Kampala and Kasese District were three research settings where the risk of tick bites was also mentioned for adults and older people, either as a statement that all people, regardless of their age, could get a tick bite, or specifically mentioning older people, as they often sit on the ground as mentioned in Kasese.

> *“And the 60years old and above, because these old people are always seated in the dirt.”* (Women’s FGD, Kasese)

Another common theme across discussions at all study sites was that domestic animals commonly harbour ticks. Participants talked about a variety of activities with domestic animals daily, including taking animals for grazing and accessing water, cleaning resting places, feeding, and milking. All activities where ticks could crawl onto the human handling the animal.

> *“I’ve never gone to a farm, and I didn’t see a tick, […] on an animal.”* (KII, Kalangala)

Participants rarely mentioned a specific sex, when discussing the daily activities with animals. But as it was mentioned in all FGDs, women, men as well as teenagers, it seems that both sexes are equally involved in activities with their animals. The women’s FGD in Kalangala stood out, as it was the only one, where sex was directly discussed and highlighted that women do look after their own animals, and are therefore in close contact with domestic animals and their ticks.

> *“It is a good thing you invited us the women because we are the ones that do that work (caring for animals). Even if it is milking. I milk my own cow. Even taking it to the bushes to eat I take it.”* (Women’s FGD, Kalangala)

Multiple reasons were discussed for why people keep animals, and which animals they keep. The main reason for most was the consumption of milk and meat from animals, discussed below. But there were also other reasons why people owned animals. One is the status in the community and linkage with power and wealth. Animal gifts to the family of the son’s bride as dowries are considered *“cultural payments”* (Women FGD Arua) and are required for weddings. Dogs and cats are kept for security reasons, pest elimination, hunting wild animals, as well as for petting in districts like Kampala, Kalangala, and Soroti.

> *“The animal is a precious bank for the communities. It is even a source of prestige because when you have animals or a herd of animals, you are a social capitalist. You have power to make decisions on affairs of the community because you will be contributing to the community. When there is a funeral, you will be looked at as a saviour.”* (KII, Arua)
>
> *“We have cats for security and [they] are believed to detect witchcraft. They are treated as family members.”* (Community leader’s FGD, Kalangala)

The reasons why people own animals inform their behaviour towards different types of animals, and how animals are kept at night. Large herds of animals are kept in kraals, which are enclosed spaces for animals to find shelter and are in proximity to people’s homes. Kraals were reported in all districts but were less common in Kasese. Animals, especially goats and chickens, were commonly reported to be kept in the main house at night in Kasese and Arua, and few times in Soroti. This practice was mentioned less frequently in the other districts. The main reason cited was the insecurity at night and the fear of theft. Additional reasons were a lack of other options due to confinement or financial means. Keeping animals in the house can mean close contact over a long period, where ticks may move from animals to humans.

> *“We also have a norm in Bakonjo that says whether a man or a woman shouldn’t miss having a goat in his or her home. […] And you don’t have where to keep it so you end up putting it in your house.”* (Community leader’s FGD, Kasese)

Only reported in Kaabong, was the practice of sleeping next to the animals in the kraals. All teenagers mentioned it, as well as a participant in the men’s FGD, to protect the animals from being stolen during raids.

> *“We sleep with animals during raids to protect them from enemies.”* (Men’s FGD, Kaabong)

Peridomestic animals were also mentioned as sources of ticks by participants. Rodents are common around houses in all study sites, can carry ticks, and get very close to humans, especially at night.

> *“You will come to notice the presence of rats when your hands and feet may be eaten up by rats at night and you will come to notice this with pain while washing your hands. When you check the hand or feet, rat teeth marks will be evident.”* (Men’s FGD, Arua)

To summarise, participants associated the risk of tick bites with particular kinds of areas, groups, and activities. Across all six settings, children were seen to be particularly vulnerable to tick bites. Overall, participants highlighted the importance of the presence of animals in the risk of tick bites.

#### Direct tick contact

Ticks are regularly removed from animals by hand or by spraying affected animals with an acaricide to kill the ticks on the animals.

> *“If they are not many, I hand pick them directly and kill them.”* (Men’s FGD, Soroti)

Regarding the risk of contact and potential transmission of CCHFV from ticks to humans, one behaviour stood out in Arua and Kaabong, which was previously highlighted in a quantitative survey but was not investigated in detail in a qualitative study before, which was the consumption of engorged ticks plucked from animals and then roasted in an open fire (8). We also observed the preparation of a tick by several herdsmen near Arua city (see **Figure 2**).

**Fig 2.**
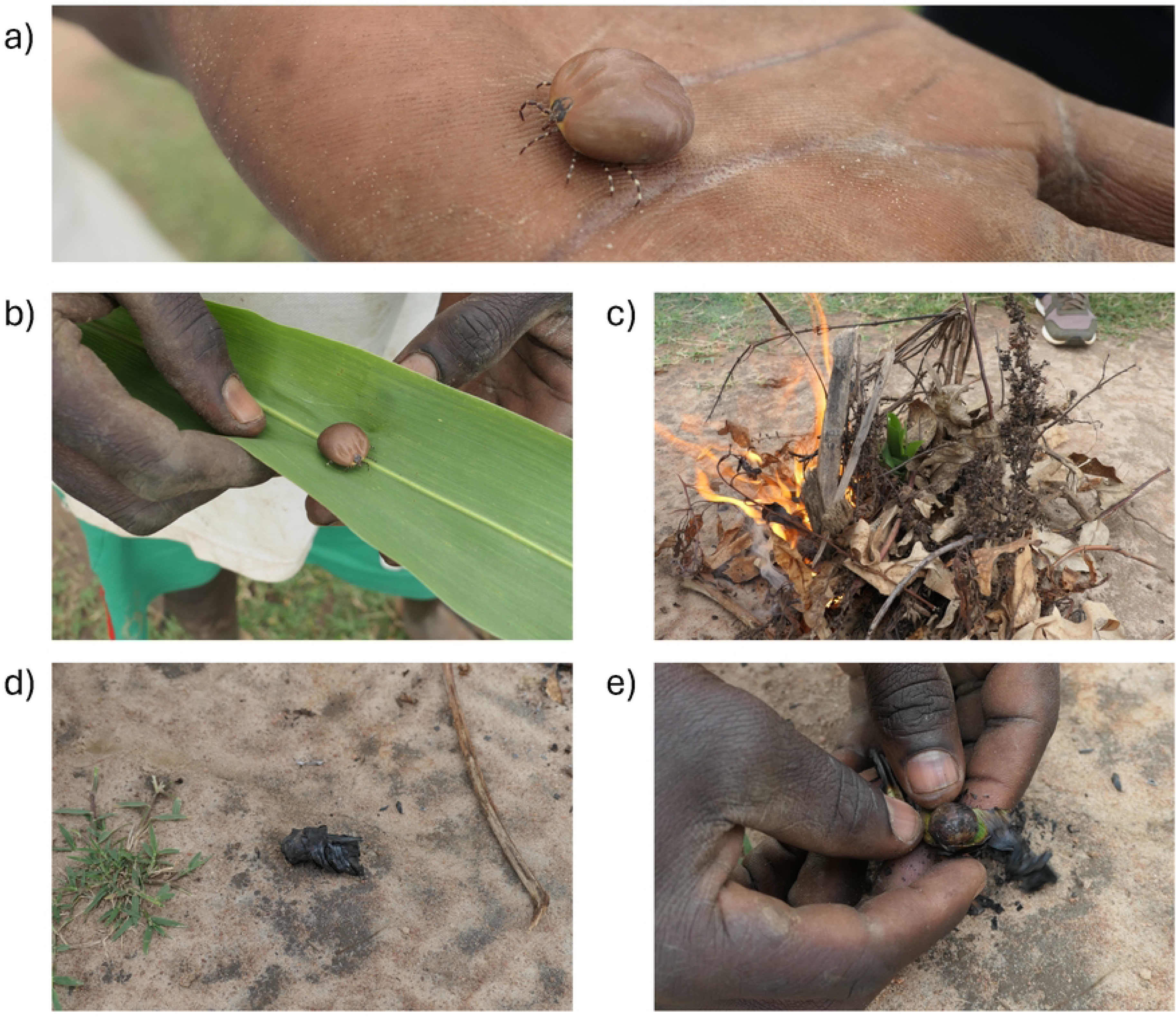
Roasting an *Amblyomma variegatum* tick in Arua. a) The female tick was extracted while fully engorged from a cow. b) The tick was next wrapped in a leaf. c) A small fire was lit, and the leaf-wrapped tick was thrown in the middle. d) The leaf-wrapped tick was removed from the fire when fully blackened, after around 5 minutes. e) The roasted tick was removed and consumed. All pictures were taken in November 2023 by MK.

Participants reported that ticks were eaten either for their protein value or that the people roasting and eating them see this as a punishment for sucking blood from their animals.

“It is very nice first of all we know that that’s the blood from the cow.” (Traditional healer, Kaabong)

“*As they eat the ticks, they make comments like ‘these are the ticks sucking blood of our animals and we must punish them*.” (Women’s FGD, Arua)

In Arua, many participants reported seeing this practice, but it was often not reported as being enacted at the time by participants in Arua. And in Kaabong, participants more often said it was a practice from the past rather than they are still doing it nowadays. However, our observation in Arua, as well as the report from the traditional healer in Kaabong, indicate that the practice is still widespread and practiced in the two districts.

### Direct contact during slaughtering

Slaughtering was another key activity providing direct contact with animals. The possible risk of direct transmission of CCHFV, through contact with the body fluids of an infected animal, was described in all districts.

> *“You cannot slaughter a cow or goat or even a chicken without getting into contact with its blood*.“ (Men’s FGD, Kampala)

Most participants in all research settings mentioned that they had slaughtered animals themselves before, or that they knew family members or community members who had carried out this activity. Slaughtering was most often reported to take place at home or at the farm, rather than at an abattoir. The remoteness of places such as the islands of Kalangala, the lack of official abattoirs, rituals for events in life like marriage, birth and death, and religious rituals are all reasons mentioned for slaughtering animals at participants’ homes. Abattoirs were only mentioned as a means to sell the meat to the public.

> *“They cannot transport the cow on the boat to bring it to the slaughterhouse and take the meat back home. So what they do is slaughter from their homes and that is it.”* (Community leader‘s FGD, Kalangala)
>
> *“What we have in Kalangala is the abattoir, but it is for only cows. For the goats, they slaughter them from anywhere, and the same applies to pigs.”* (Community leader’s FGD, Kalangala)
>
> *“Like when the rain takes long to fall, the elders go to the shrine, and they pick a cow and slaughter for the gods and also take it when it’s fresh.”* (Teenager’s FGD, Kaabong)

When animals are slaughtered privately, it is unlikely that there are any veterinary personnel who inspect the animal pre- and post-mortem. This may then lead to or enable the slaughtering of sick animals or butchering and consuming animals which died a natural death. Participants in all districts except Kampala and Arua mentioned the slaughtering of sick and dead animals for consumption. Various reasons were mentioned, including the commercial value of the animal, its nutritional value, or ignorance of the possibility of transmitting diseases.

> *“Facilitator: You don’t throw the meat away [if the animal died naturally]? Participant: No we don’t, we eat the meat.”* (Herdsman, Kaabong)
>
> *“If my cow is sick, do you think I will bring it here [the abattoir]? We just slaughter it from the farm, share the meat and call it a day.”* (Community leader’s FGD, Kalangala)

### Animal product consumption

Consuming meat, blood and other animal products from previously infected animals may be a lower risk transmission route for CCHFV based on few reports in the literature. However, a few practices stood out during the interviews and discussions, highlighting the possibility of transmission of CCHFV infection, through consuming sick animals or wild animals, as well as eating uncooked, or only partially cooked products.

As described earlier, participants described multiple reasons why sick animals were slaughtered and later consumed, particularly the value of the meat, which trumped possible infection risks through consumption.

> *“You cannot throw away meat. That is meat. Meat remains meat. I cannot throw my cow.”* (Women’s FGD, Kasese)

Wild animals, including antelopes, wild birds, wild large rats, and others, were described as being hunted by members of the communities. These were reported to be infested with ticks and may be associated with a risk of transmission of known and unknown infections. These species have an unknown risk of harbouring CCHFV infection. Soroti, a participant mentioned that boys in the community are hunting birds.

> *“There is a tendency of young boys having catapults. They were looking after cows at the same time hunting birds down.”* (KII, Soroti)

Reports of meat being eaten partially cooked or even raw were common in several districts. Consuming partially cooked meat was reported when animals might be roasted as a whole and middle parts might not be completely cooked, when animals were roasted in the forests or shrines for ritual, or when there was a sense of urgency.

> *“[When] like around 20 people are waiting so you have to be fast enough to meet the demand [of roasted pork at a place of excitement like a bar]”* (Community leader’s FGD, Kalangala)

In addition to eating meat, consuming animal blood was commonly reported by multiple participants in Kasese, Kaabong, Arua and Soroti Districts. However, there were differences in the preparation and the means of consumption. In Kasese, participants discussed rearing guinea pigs, which are slaughtered for the primary purpose of drinking their fresh, uncooked blood to fight anaemia. In Arua, a blood meal is cooked in advance of consumption, but holds a strong cultural meaning, as mentioned by multiple participants. Blood is consumed raw in Kaabong, often directly still warm from a prick in the animal’s neck. It was also seen as a nutritional meal, similar to reports in Kasese.

> *“You just cut it [the guinea pig] and take its fresh blood fighting anaemia.”* (Community leader’s FGD, Kasese)”
>
> *“In addition [to preparing the blood], they also squeeze the faeces (fresh dung) which is still in the animal’s stomach and mix with blood as they cook.”* (Community leader’s FGD, Arua)

Blood was also frequently mentioned during rituals, such as sacrifices, healing, births, funerals, and weddings. It may be poured or smeared on a person or people bathe in it.

> *“When you go to a shrine, you are sent for a goat. The goat is cut, and you are bathed with the blood.”* (Community leader’s FGD, Kalangala)

Overall, a range of potential risk factors were mentioned relating to tick bites and tick presence, consumption of ticks, slaughtering and eating meat and other animal products and direct contact with healthy and sick animals and carcasses. Some practices varied strongly by district while others were widespread within most districts. This suggests that a one-size-fits-all approach may be less effective than developing strategies that respond to the particular practices of a community.

## Discussion

In this study, we used FGDs and KIIs to explore people’s views on human-animal-tick interactions in six distinct districts of Uganda. We aimed to understand the transmission risk of CCHFV, primarily through tick bites and tick consumption, direct contact with live animals and during slaughtering or consumption of animal products.

Participants described differences in perceived tick burden that correlated with reported tick-bite prevalence across all FGDs. The highest reported tick abundances occurred in the northern and western districts: Kaabong and Kasese. Consumption of engorged ticks was reported in Kaabong and in Arua District. Slaughtering at home and slaughtering sick animals were very common in all districts. The consumption of raw blood was limited to Kaabong and Kasese districts.

In our study, the highest reports for both seeing ticks and receiving tick bites were in Soroti and Kasese districts, and the lowest in Kampala and Kalangala districts. This link between human tick bites and the abundance of ticks in the environment has been investigated in orienteers in Scotland, where they found a strong correlation with reported tick bites and a questing tick survey (ticks ready to bite a passing animal or human) (36). This study investigated a very specific activity only in one region of the world, but highlights that the perception of our study participants that the tick burden in the environment and tick bites can correlate. Similar studies in Uganda could show this correlation in quantitative ways, as well as look at more relatable activities to the Ugandan population, like grazing animals or working in a field.

Reports about tick observations were mainly mentioned in relation to animal contact or being in areas where animals eat, rest, or drink. Domestic and wild animals are commonly infested with ticks (37–39) and are known to be asymptomatic reservoirs for CCHFV (1,40). That participants mention the connection between tick reports and animals and the environments where they spend time is therefore consistent with general knowledge. As the ticks bite the animals to survive and moult to their next life stages, they can become infected with CCHFV and can spread it during their next bite. Therefore, these ticks pose an exposure risk of infection to humans.

Activities with domestic animals, such as feeding, cleaning, or bringing them for grazing were not distinctly separated by sex in our study population. It was not probed specifically by our research team but also did not come up in most conversations except the women’s FGD in Kalangala. We did also not separate conversations around animal husbandry and care between small and large animals and animal herds, which might have caused different discussions. Agaba et al. (2025) discussed clear gender roles regarding small animal groups at the homestead and contact with larger herds for commercial purposes (21). However, if the exposure risk to CCHFV is not proven to be higher or lower within the different activities, both sexes are at risk while handling animals, and preventive measurements should be planned for both activities.

Rather than gender differences dominating perceived tick-bite risk, our study found that participants identified children as the highest-risk group of tick bites. However, there is a lack of scientific literature in Uganda to identify the actual specific risk of tick bites by age group, including with respect to CCHFV exposure. The most frequent age group among reported severe CCHF cases in Uganda is 21 to 30 years (7). However, children might generally present with a milder course of CCHF (41) and wouldn’t be included in the above statistic. Investigating other tick-borne diseases, within the Lyme disease surveillance in the USA, the highest risk was reported for children up to the age of 14 (42), and a similar study in Canada reported that children (5-9 years) and older adults (50-79 years) had the highest incidence rates for Lyme disease (43). We hypothesise that if tick bites and exposure to CCHFV are investigated in Uganda, the pattern will be similar to that observed in other studies, with milder disease in children, but a higher incidence or force of infection compared to adults.

Occupational risk related to daily handling of animals was evident in the conversations with participants, and several occupations were mentioned as likely to put people at higher risk of tick bites and potential CCHFV exposure, including herdsmen, abattoir workers, farmers, hunters and veterinary personnel. Other studies have shown that tick bites are common in certain professions, for example, in forest workers and farmers in Germany (44). However, such occupations are highly context-specific and differ by country and even region, for example, with large forests present in Western Uganda and large grazing areas in Central Uganda, leading to different occupations. Furthermore, tick abundance, tick species, and tick bite prevention are context-specific, which is why our reports from our study sites highlight Ugandan-specific occupations to be further studied for the risk of tick bites.

The perceived risk of tick bites from ticks that have come off wild animals in national parks is supported by multiple studies in Uganda and other African countries. Ticks have been seen and collected from chimpanzees in Kibale forest of Uganda (45,46); in a review including studies from 17 African countries, it is reported that ticks have been collected from 35 different wild animal species and harboured 34 infectious agents, including CCHFV (47); and several wild animal species are well-reported to be exposed to CCHFV and have high seroprevalences in studied populations (48,49), indicating that ticks feed on wild animals. These studies support the communities’ perceived risk of tick bites from ticks on wild animals and further highlight the risk of CCHFV exposure from these.

Eating engorged ticks in Uganda was first mentioned in scientific literature by Atim et al. (2022) and was significantly positively correlated with higher exposure rates to CCHFV in Arua district (8). We have, for the first time, recorded that not only in Arua but also in Kaabong, people consume engorged ticks, which likely increases their risk of CCHFV exposure. Eating engorged ticks has also been reported in other countries and is not unique to Uganda. A recent study in Cameroon found that 3% of interviewees reported having eaten ticks (12). A missing awareness of this practice by researchers, and a reluctance to report a stigmatised practice could mean that it might be a more common potential CCHFV exposure risk than current data suggests. Future studies on the biological significance of this transmission risk are needed, and tick ingestion should be included in all studies examining the risk of tick-borne virus exposure.

While all participants mentioned that ticks were roasted before consumption, there is a chance that ticks might be eaten before virus particles are completely denatured, as efficient heating is required to denature the viral particle (50). Conversely, it is possible that sometimes viral proteins are sufficiently denatured and could hypothetically act as a mucosal vaccine against CCHFV infection. This, alongside exposure to other nairoviruses, known to occur commonly in ticks in Uganda, might in fact provide a protective effect for severe disease and could therefore be beneficial for the communities who were or are still consuming engorged ticks, as reported in our qualitative study.

A very high-risk route of infection is likely to occur while plucking ticks from animals with bare hands. Blood could enter through wounds or micro-cuts/abrasions, allowing the virus to cross the skin barrier to potentially infect that person. Direct transmission from CCHFV patients’ blood or body fluids in hospital settings to healthcare workers through skin contact has been well described, highlighting that this is likely a potential risk activity (51).

Another route of direct exposure to CCHFV is expected when slaughtering infected animals or consuming infectious animal products. CCHFV can enter through microcuts in the skin or mucosa when a virus particle from blood or body fluids comes into contact with the individual (2). Slaughtering may be associated with particularly high exposure, as evidenced by the high risk of CCHFV exposure among abattoir workers (52,53). The common reports of slaughtering at home by multiple participants open the possibility of higher exposure in Ugandan communities, as individuals might have less training, expertise and experience than professional abattoir workers. Evidently, for home slaughter, there would not be a supervising veterinary officer present to check on the animals pre- and post-slaughtering, to identify sickness and prohibit activities if necessary. This was one of the reasons mentioned by the participants regarding why they slaughtered from home, to avoid such controls and be able to slaughter and eat sick animals, and not to lose precious meat sources.

Infections through food intake have also been reported from uncooked and infectious meat (54,55). It is not only in our study that it is reported that sick animals are slaughtered and eaten, which could increase the risk of CCHFV exposure. For example, in a study in Kenya, Cook et al (2017) reported that 9% of slaughterhouses slaughtered and sold sick animals (56), and in a Nigerian study, Sylvia et al (2024) reported 55% of participants believed that diseases could not be contracted from eating sick animals (57). These suggest a widespread behaviour across CCHFV-endemic countries in Africa, which could be targeted in high-risk areas, during a wider outbreak, or special celebrations like Eid al-Adha, Christmas or Easter, to reduce infections. For example, a 14-day quarantine and acaricide treatment to control ticks in animals before slaughter could reduce the risk of transmission from animals to humans (58).

This study was designed to capture a variety of behaviours and differences in human, animal, and tick interactions across Uganda, using K-prototype analysis initially to select heterogeneous districts before focusing on six distinct areas for the study. Several high-risk activities were highlighted, and the heterogeneity of the FGDs and KIIs findings in the different districts supports the care taken to locate geographically and culturally different areas of the country. However, while these findings are important, the study could not fully represent the whole of Uganda and wider endemic areas for CCHFV. Recruitment was opportunistic within selected districts, based on the availability of participants and existing relationships with community leaders. In the field of local ecological knowledge (LEK), researchers have discussed identifying experts within communities and state that rigorous reporting and a more systematic approach, such as identification by peers or systematic surveys, could help document knowledge more thoroughly (59). For future work on CCHFV, this would especially apply to methods of controlling tick burden in the communities, as these data would benefit from more rigorous documentation.

While the selection of social scientists who spoke the local languages fluently in this study was a definite advantage in the study, the use of multiple different researchers conducting KIIs and FGDs may have somewhat biased reporting on differences when questions were asked in a slightly different style. However, the interviewers all received training in the topic guide, and differences were minimised where possible.

### Conclusion

This study highlighted extremely close human-animal-tick contact across six diverse Ugandan districts, with practices such as animal cohabitation and slaughtering of sick or dead animals, alongside daily tick exposure for some participants, which could all potentially contribute to elevated CCHFV exposure and transmission risk. Specific behaviours, including roasting engorged ticks, private slaughtering and eating of sick animals, and consuming fresh animal blood, underscore the need to quantify the frequency of these practices further and assess the actual level of risk they may pose and how this might inform behavioural changes on a day-to-day basis, as well as for potential outbreak scenarios. Such insights are essential for informing contextually appropriate public-health strategies. These findings also reinforce the critical importance of integrating qualitative perspectives into epidemiological research.

## Data Availability

The data will be made available in fully anonymised form following publication of the related manuscripts.

## Acknowledgements

We would like to acknowledge Prof. Janet Seeley and Dr Suzan Trienekens for their support and advice during the inception of the project, as well as Prof. Kimberly Fornace and Emilia Johnson, for their support with the spatial analysis and district selection process. We want to thank the administrative personnel from both the Medical Research Council–University of Glasgow Centre for Virus Research and the Uganda Virus Research Institute for their help with finances and logistics. We would also like to acknowledge the Village Health Team members and the local council leader for the mobilisation of participants. We also thank the district officials across the six districts for their support with administrative clearance and for introducing the study team to the village health teams during the study. In a special way, we would like to thank all participants for taking the time to take part in the study.

